# Informative Frailty Indices from Binarized Biomarkers

**DOI:** 10.1101/2020.01.07.20016816

**Authors:** Garrett Stubbings, Spencer Farrell, Arnold Mitnitski, Kenneth Rockwood, Andrew Rutenberg

## Abstract

Frailty indices (FI) based on continuous valued health data, such as obtained from blood and urine tests, have been shown to be predictive of adverse health outcomes. However, creating FI from such biomarker data requires a binarization treatment that is difficult to standardize across studies. In this work, we explore a “quantile” methodology for the generic treatment of biomarker data that allows us to construct an FI without preexisting medical knowledge (i.e. risk thresholds) of the included biomarkers. We show that our quantile approach performs as well as, or even slightly better than, established methods for the National Health and Nutrition Examination Survey (NHANES) and the Canadian Study of Health and Aging (CSHA) data sets. Furthermore, we show that our approach is robust to cohort effects within studies as compared to other data-based methods. The success of our binarization approaches provides insight into the robustness of the FI as a health measure, the upper limits of the FI observed in various data sets, and highlights general difficulties in obtaining absolute scales for comparing FI between studies.

## Introduction

Poor health is often associated with aging, a decrease in functional capacity, and an increased susceptibility to illness and injury. While chronological age is a convenient proxy for aging, it cannot capture individual variability of health at a given age. The frailty index (FI) is a well-tested way of incorporating large and varied aspects of health and function that can be easily used to differentiate between individuals of the same age. Defined as the fraction of selected health attributes that are in an unhealthy state (called deficits), the FI has been shown to be a robust measure of individual health over the aging process (Mitnitski et al. 2001, Searle et al. 2008). The FI is observed to increase with age and the distribution of FI on a population level broadens with increasing age, describing the heterogeneity of aging (Gu et al. 2009). The FI is predictive of mortality and of other adverse health outcomes (Rockwood, Song, MacKnight, Bergman, Hogan, McDowell & Mitnitski 2006, Evans et al. 2014).

The health attributes considered in the FI are typically clinically observable or self-reported, such as disabilities in activities of daily living or physical or cognitive impairments (Searle et al. 2008). Alternatively, standard laboratory measurements such as blood and urine biomarkers (Blodgett et al. 2017, Mitnitski et al. 2015, Howlett et al. 2014) as well as biomarkers of cellular senescence and oxidative stress (Mitnitski et al. 2015) can be used to create a laboratory-test based FI known as FI-Lab. Cutpoints are used to binarize the quantitative biomarker measurements into deficits so that they can be naturally included in an FI. Normal reference ranges based on diagnostic or therapeutic utility (McPherson 2017) are commonly used as cutpoints.

Since the FI is an aggregate measure and is not used for the diagnosis or treatment of specific conditions, standard cutpoints are not necessarily best suited to its role of predicting risks. Furthermore, standard cutpoints are often not available for emerging biomarker measurements such as in epigenetic, proteomic, metabolomic, or other high-throughput “omics” approaches. Alternative “data-based” methods obtain cutpoints from the available data under consideration. Both normal reference ranges (Blodgett et al. 2017, Howlett et al. 2014) and data-based methods (Mitnitski et al. 2015) can and have been used to create an effective FI-Lab.

One data-based method of biomarker binarization is to select cutpoints to maximize some predictive aspect of the post-binarized biomarker. For example, cut-points can be selected to maximize the difference between survival curves of people that are on either side of the cutpoint for each biomarker (Mitnitski et al. 2015). Equivalently, other predictive measures such as receiver operator characteristics (ROC) performance or mutual information (Farrell et al. 2016) could be used with respect to a particular outcome such as mortality within 5-years to generate “optimal” cutpoints. While attractive in principle, such individual biomarker optimization approaches run the risk of creating FI that are overly specific to the study cohort and not generally applicable for other cohorts.

Another popular data-based method for binarizing continuous-valued data is to select cutpoints based on the quantile of the population. This approach is used in both the Fried frailty phenotype (Fried et al. 2001) (with quintiles) and in the exploration of the allostatic load theory of physiological disregulation (Seplaki et al. 2005, Juster et al. 2010) (with quartiles). Here, a risk direction is chosen for each biomarker, e.g. by how the biomarker changes with age, and the cutpoint is selected for each biomarker by the quantile of that biomarker – i.e. the fraction of the population that has values of the biomarker above the cutpoint. This approach should be less susceptible to overfitting, since the quantile is chosen globally for all biomarkers rather than individually for each biomarker. Nevertheless, it raises the question of how to choose the best quantile and of how sensitively the results depend upon the quantile chosen. Investigation of allostatic load (Seplaki et al. 2005) found that deciles and quartiles behaved similarly, implying that the quantile approach may be robust with respect to choice of quantile. Nevertheless, no systematic investigation of the quantile approach in the context of the FI has been done before.

A systematic investigation of data-based approaches for the binarization of continuous-valued biomarkers used in the evaluation of the FI can explore the questions of overfitting due to optimization raised above. At the same time, we can examine the robustness (or insensitivity) of the FI as a predictive measure of health outcomes or mortality and the robustness of the FI maximum seen in observational studies of aging (Searle et al. 2008, Mitnitski et al. 2015), with respect to the details of any binarization approach. Robust and validated data-based approaches to binarization will facilitate the future development of FI for high-throughput ‘omics data and for more model organisms of aging.

Here, we examine the effectiveness of data-based binarization schemes for building the FI from biomarker data. We use both the NHANES and CSHA data sets to check whether cohort effects are large; we find that they are not. We examine overfitting effects with cross-validation, and find that they are present when optimal cutpoints are chosen for each biomarker – but that they are small when global cutpoints are chosen for all biomarkers. We compare the predictive performance of data-based schemes against earlier published results, and find that the data-based schemes have comparable or slightly higher predictive value than the established FI with respect to predicting mortality and clinical deficits. Overall, we find that a generic quantile data-based binarization approach performs well.

A key characteristic of the FI is the relatively insensitivity (Searle et al. 2008, Mitnitski et al. 2015) to the particular choice of deficits. We show that this also holds for choosing cutpoints for FI-Lab, and we find that a broad range of cutpoints exist where the quantile binarized FI-Lab is effective. This demonstrates both the universality of the FI and the generality of our method of choosing cutpoints. Nevertheless, we identify the best range of quantiles to use and we find that they overlap with the quintiles used in the Fried frailty phenotype (Fried et al. 2001). Furthermore, many aspects of the FI calculated at these quantiles such as maximum, minimum and overall distribution of FI in the population overlap with results from previous FI-Lab studies.

## Methods

### Data, evaluation, and cross-validation

The data used in this study are from the National Health and Nutrition Examination Study (NHANES) (Centers for Disease Control and Prevention National Center for Health Statistics Updated 2014) and the Canadian Study of Health and Aging (CSHA) (Canadian Study of Health and Aging Working Group 1994). The NHANES data set consists of the 8881 individuals from the NHANES study with data for at least 11 of the 16 available biomarkers. This sample has an age range of 20 to 85. The data used from the CSHA study has 973 individuals aged 65+ for which data is available for at least 16 of the 22 biomarkers. Age distributions for these data sets are shown in supplemental Fig. S1, which highlights the smaller and older cohort of the CSHA study.

Both of these data sets have previously been used to construct FI-Lab. Blodgett *et. al* (Blodgett et al. 2017) considered the NHANES data set, while Howlett *et. al* (Howlett et al. 2014) considered the CSHA. We will compare our results with both of these in this paper. Since a much larger sample size and a much larger range of ages are available, we focus on the NHANES data set. However, major results will be also validated in the CSHA data set. Both studies’ FI-Lab consist of many shared deficits and cutpoints, so the differences in FI between the data sets are likely due to cohort effects. These two data sets have very different cohorts, so by applying our methods to both we test the generalizability of our approach.

The NHANES and CSHA cohorts differ in more than just age. In supplemental Fig. S2 we show the distribution of FI-Lab for the CSHA cohort (white bars) together with a resampled NHANES cohort with the same (65-85 years) age distribution as the CSHA (blue bars). We see that the NHANES cohort has a significantly lower FI-Lab at the same age, i.e. it represents somewhat healthier individuals. This could be due to a large portion of the CSHA population being comprised of institutionalized individuals (Howlett et al. 2014).

The purpose of binarizing data is to construct an FI. The FI is intended to be an inclusive and general indicator of individual health; it has been shown to correlate well with mortality (Kojima et al. 2018) but also with institutionalization (Rockwood, Mitnitski, Song, Steen & Skoog 2006), postoperative complications (Velanovich et al. 2013), dementia (Song et al. 2014), recovery time in hospital (Hatheway et al. 2017), and other adverse health outcomes (Blodgett et al. 2016). Accordingly, we compare our newly constructed FI with the existing FI-Lab in their ability to predict 5 year mortality as well as by their ability to predict clinical outcomes from laboratory data. To evaluate prediction, we use the standard area under the curve (AUC) of the ROC curve. We obtain similar results using mutual information (Farrell et al. 2016), as illustrated in supplemental Fig. S3. We also check that our new FI behave similarly to the previously published FI-Lab, with respect the clinical FI, with respect to their distributions, and with respect the maximal observed FI in the population.

Each new FI is tested using cross-validation. Cut-points are generated using a random half of the population, then those cutpoints are applied to the other half and the resulting FI are evaluated. This is repeated 100 times. Cross-validation allows us to characterize any over-fitting of cutpoints.

### Quantile-based cutpoints

We transform the biomarker data to a dimensionless form using quantiles. For each individual subject, each biomarker *i* is transformed to the proportion *x*_*i*_ of the population that has “less risky” values. This is illustrated in Fig. 1 for systolic blood pressure. If an individual has a value of 140 mmHg, which places them at the upper quintile of risk for blood pressure, their systolic blood pressure score is transformed to *x* = 0.8, corresponding to having a higher systolic blood pressure than 80% of the population.

**Fig. 1.**
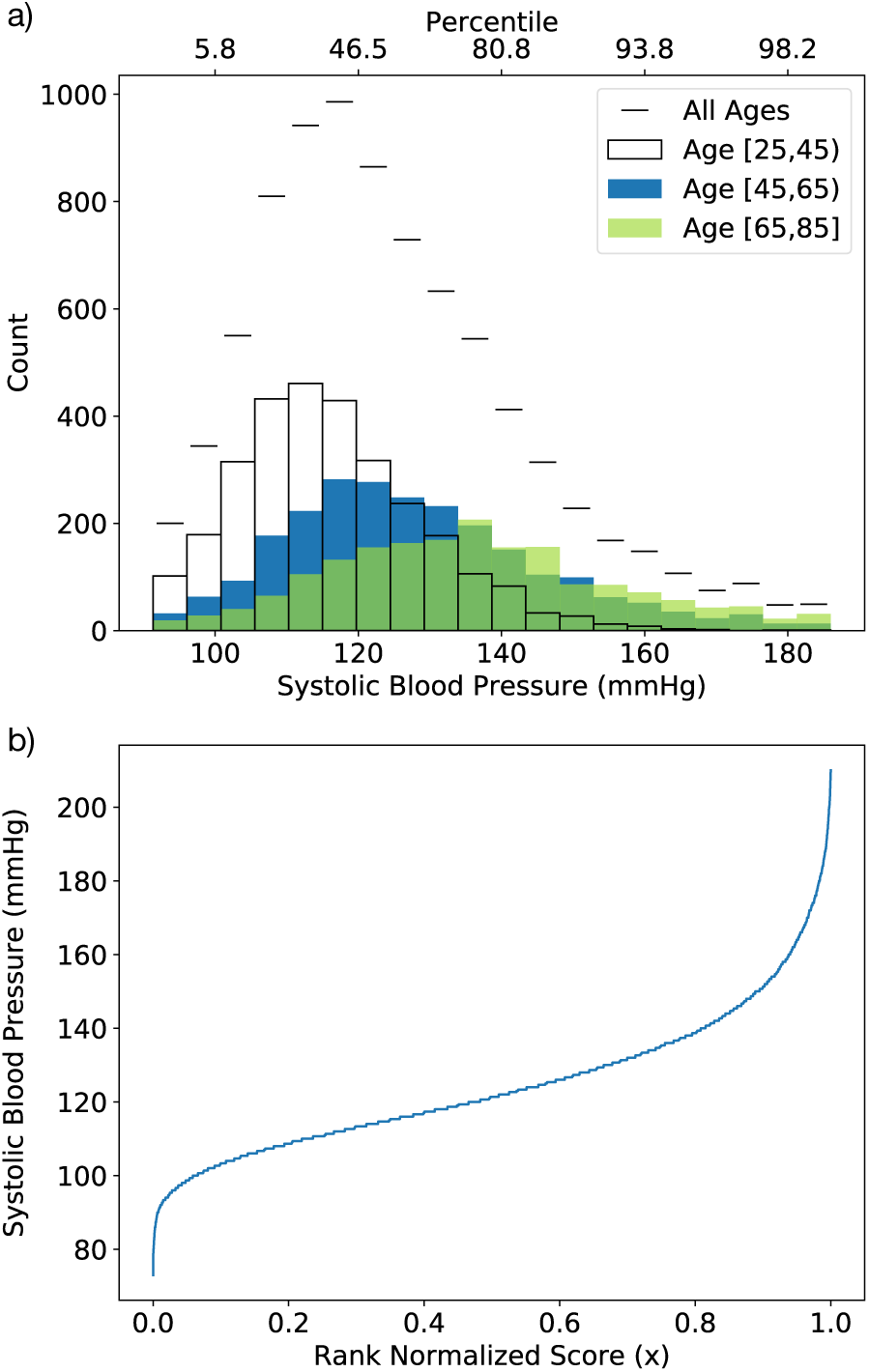
a) The distributions of systolic blood pressure measurements in the NHANES cohort (Centers for Disease Control and Prevention National Center for Health Statistics Updated 2014). Short horizontal lines indicate the whole population distribution, while unfilled, orange, and blue bars show the youngest [25, 45), middle [45, 65), and oldest [65, 85] age groups respectively (in years). The trend during aging is an upward shift. b) The rank normalized score *x* vs the corresponding systolic blood pressures. The median corresponds to *x* = 0.5. For this and other measures, the nonlinear mapping between *x* and corresponding value is always monotonic – but is either increasing or decreasing depending on the direction of risk.

Quantiles are implemented on a population scale by performing a rank normalization of the data, where each biomarker is sorted in ascending risk, then the ranks (position in the sorted list) are divided by the number of individuals. The rank normalized values *x*_*i*_ are given by

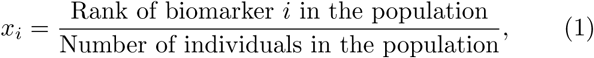

and so *x*_*i*_ ∈ [0, 1].

Implementing binarization is straightforward using these quantiles. We apply a global cutpoint (GCP) as a threshold value of the rank normalized values, *X*_*GCP*_, applied identically across all biomarkers. We build the resulting FI as the average over these binarized deficits,

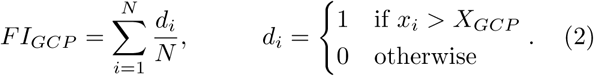

For each biomarker, a deficit *d*_*i*_ = 1 is assigned when *x*_*i*_ passes the threshold in the direction of risk.

### Direction of risk

We determine a direction of risk for each biomarker, before applying quantile-based cutpoints. We then binarize with respect to the at-risk direction, as discussed above. We do not assert that biomarkers only have one direction of risk, but we do find that most biomarkers have one direction that is most often explored by the population, and so we assume this is the dominate direction of risk during aging. This is illustrated in the supplemental Fig. S4.

We prefer a mortality-free approach to determining direction of risk to avoid potential over-fitting. We simply use the aging trends of the biomarkers to determine the risk direction. The relation between age and each biomarker is determined by the sign of Spearman’s rank correlation. A positive value indicates the risk direction is towards large values of the biomarker, a negative value indicates risk towards small values. This method is effective at determining risk directions if the population has a reasonably large distribution of ages. Aging trends effectively classify risk direction in both the CSHA (ages 65-104 years) and NHANES (ages 20-85) data sets. However, we restrict the age range for calculating risk directions to ages 35+ to calculate relations based on normal aging behaviour.

Another method of determining risk direction is to use mortality data, or some other adverse health outcome. For each biomarker ROC curves can be generated with respect to the binary outcome (e.g. 5 year mortality) and an AUC can be calculated. An AUC above 0.5 indicates the primary risk direction is towards high values, an AUC below 0.5 indicates risk towards the low end. Equivalently, one could do a logistic regression of the biomarker against an adverse outcome and use the sign of the beta value (positive beta would indicate risk towards high values). This type of approach ensures that the risk directions generate the best FI for predicting that outcome, but they are potentially over-fit to that outcome. We find that risk directions from mortality data are predominantly the same as the aging trend directions. The predictive AUC of the resulting FI with respect to 5 year mortality is also essentially the same as with aging trends, as shown in supplemental Fig. S5.

We have also considered a simple approach for two-sided cutpoints. For simplicity, we consider symmetric cutpoints with both *x*_*i*_ *> X*_*GCP*_ and *x*_*i*_ *<* 1 − *X*_*GCP*_ assigned as deficits with *d*_*i*_ = 1. The predictive AUC of the resulting FI is significantly worse than the one-sided approach, as indicated by the supplemental Fig. S6. Accordingly, we focus on one-sided cutpoints in this paper.

### Optimally predictive binarization

In addition to quantile binarization, we also compare with two different FI created with cutpoints selected for optimal prediction with respect to mortality. For both, additional details are provided in the supplemental information.

The first, *FI*_*logrank*_, based on the separation of survival curves, has been used to create an FI-Lab (Mitnitski et al. 2015). For each biomarker, the cutpoint is found that maximizes the significance of separation between survival curves of individuals with and without the deficit by minimizing the p-value from a logrank test (Mantel 1966).

The second method for generating optimal cutpoints is based on information theory. *FI*_*info*_ uses cutpoints selected for the highest possible mutual information with respect to mortality at 5 years. In a manner similar to *FI*_*logrank*_ every possible cutpoint is tested for every biomarker and the cutpoints which maximize the mutual information with respect to mortality are selected.

## Results

To evaluate the various data-based approaches to binarization, we have calculated the AUC with respect to five-year mortality for both the NHANES and CSHA cohorts. The results are shown in Fig. 2. The performance of all measures was qualitatively similar for both the NHANES and CSHA data sets, though due to the smaller cohort the CSHA data showed greater variability in cross validation.

**Fig. 2.**
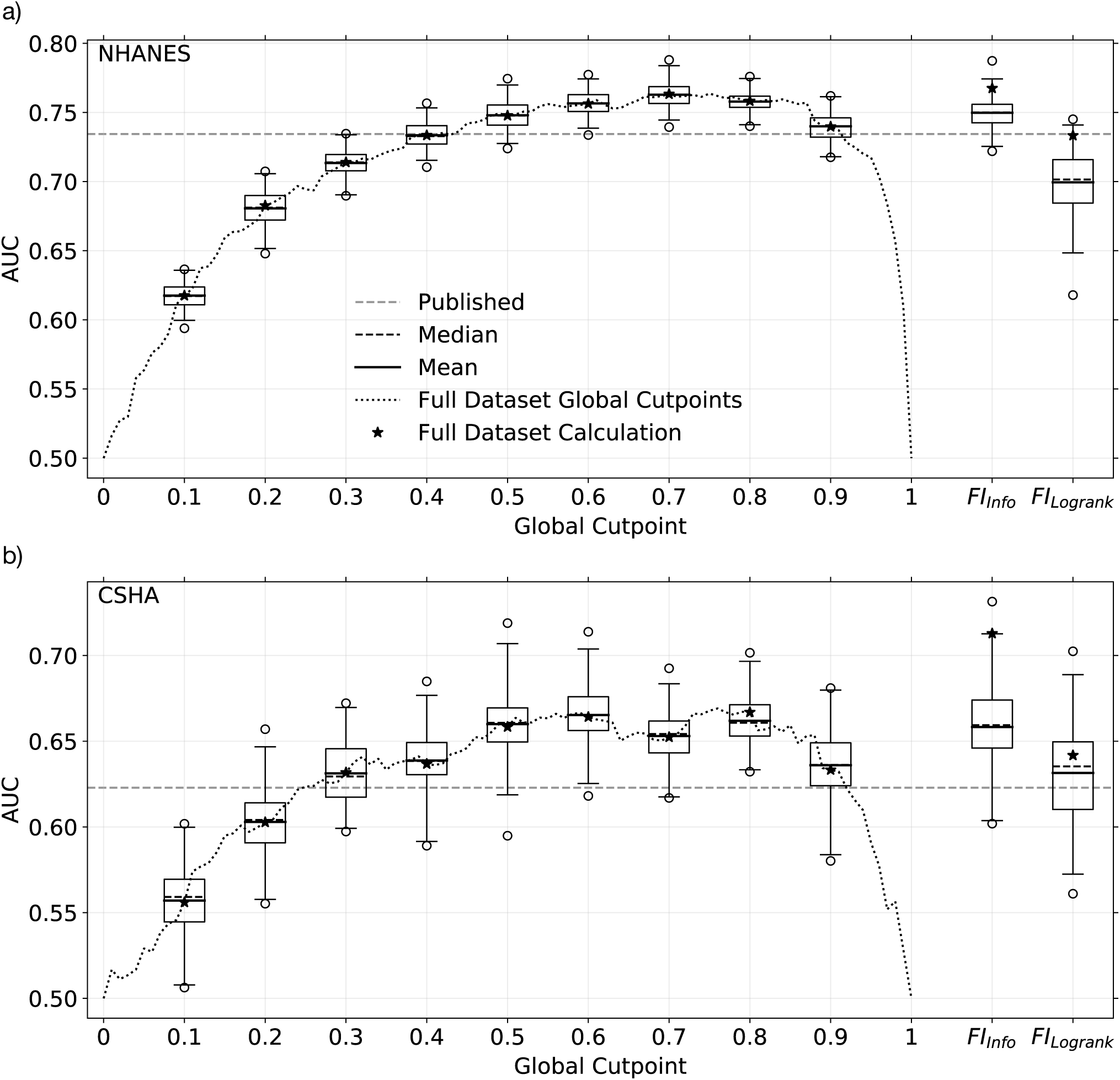
Cross-validated AUC of different data-based FI with respect to 5 year mortality for the a) NHANES and b) CSHA cohorts. The horizontal dashed line shows the AUC of the published FI (Blodgett et al. 2017, Howlett et al. 2014). The dotted line indicates the AUC of the quantile-based *FI*_*GCP*_ vs the global cutpoint *X*_*GCP*_. Box and whisker plots display the data from cross-validation: the boxes represent the upper and lower quartiles, the whiskers go to the 99^*th*^ and 1^*st*^ percentiles, and the circles are remaining outliers. The short dashed line within each box is the median, the solid line the mean, and the star is the AUC for the full data set without cross validation. The rightmost three columns, as indicated, show the AUC for *FI*_*info*_ constructed from maximum information cutpoints and *FI*_*logrank*_ constructed from logrank minimum p-value cutpoints.

*FI*_*GCP*_, assembled from quantile based global cut-points, performed well. For all tested values of *X*_*GCP*_ the cross-validated and full dataset results agree, indicating minimal overfitting. For the extreme values of *X*_*GCP*_ equal to 0 (where all biomarkers are at risk) or 1 (where none are), there is no predictive value of *FI*_*GCP*_ and the AUC is equal to 0.5 – as expected. Between these extremes, we see a broad maximum of the AUC. Indeed, for global cutpoints between 0.5 − 0.9 the *FI*_*GCP*_ slightly outperforms the published FI-Lab for both the NHANES and CSHA datasets.

The binarization approaches to maximize the mortality prediction for the full datasets gave comparable AUC values, as indicated by the columns to the right in Fig. 2. However, cross-validation of *FI*_*info*_ and *FI*_*logrank*_ showed significantly lower AUC and so indicates that these methods are prone to overfitting. While the cross-validated *FI*_*info*_, using maximum information cutpoints, appears to perform as well as *FI*_*GCP*_ – the cross-validated *FI*_*logrank*_ does not. We have shown that using individual cutpoints optimized for each biomarker to predict mortality leads to an *FI* that is prone to overfitting. Using such an optimized approach without cross-validation should be avoided. We focus on the quantile cutpoints for the remainder of this paper.

We were surprised that the quantile-based cutpoints performed similarly well for both the NHANES and CSHA datasets, despite their significantly different age, health, and cohort sizes. Since quantile-based cutpoints are extracted from the cohorts being characterized, we wanted to investigate cohort effects more directly. Since the NHANES dataset has a large population and a large range of ages, we obtained quantile-based cutpoints from sub-cohorts of NHANES for young (25-45), middle (45-65), or old (65-85) age groups. Remarkably, the cutpoints obtained from any one sub-cohort worked reasonably well applied to any other cohort. However, the range of *X*_*GCP*_ for best prediction decreased and shifted closer to 1 as shown in supplemental Fig. S7. This supports our observation that cohort effects are not large with quantile-based cutpoints.

A crucial test of FI-Lab behavior is how well it corresponds to an established FI-Clin. The coloured lines in Fig. 3 shows average *FI*_*GCP*_ values binned by their corresponding FI-Clin values. For intermediate values of *X*_*GCP*_, we see that *FI*_*GCP*_ is monotonically increasing with FI-Clin. Indeed, the published FI-Lab appears to correspond to *X*_*GCP*_ values between 0.8 and 0.9 – where the *FI*_*GCP*_ also performs well with respect to both AUC and cohort effects. Conversely, for much larger or smaller values of *X*_*GCP*_, where the AUC is significantly worse than for the published FI-Lab, we see that the *FI*_*GCP*_ is not strongly dependent on FI-Clin or even becomes non-monotonic.

**Fig. 3.**
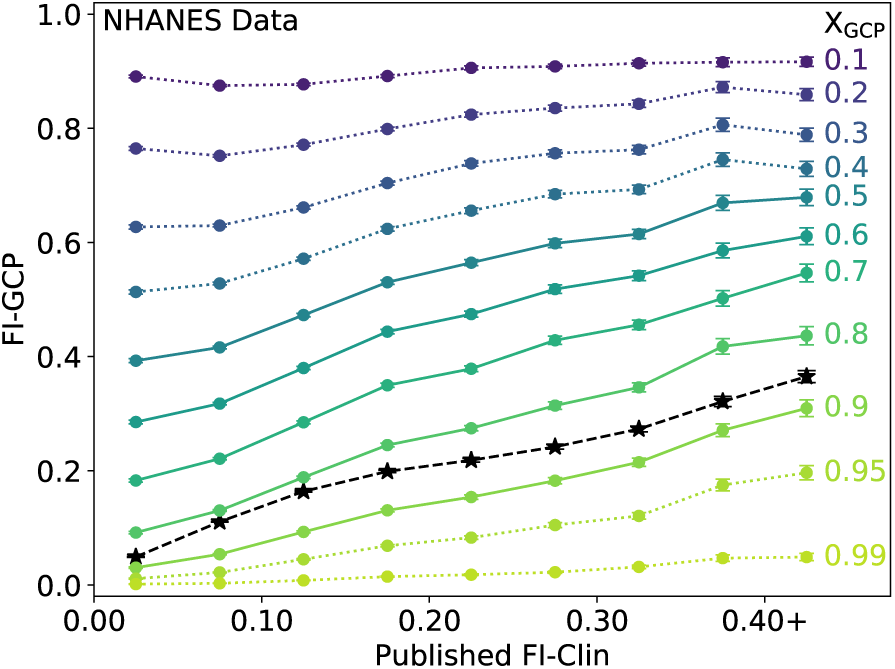
Average *FI*_*GCP*_ vs published FI-Clin for the NHANES dataset, for a variety of global cutpoints *X*_*GCP*_ as indicated by the coloured numbers at the right of each coloured line. The coloured markers indicate the middle of the bins used for averaging. The black dashed lines with stars show the published FI-Lab (Blodgett et al. 2017). The *FI*_*GCP*_ lines are dotted when their AUC from Fig. 2 is below the published value, while they are solid when it is above.

We test the versatility of the FI by its ability to predict outcomes other than mortality. In Fig. 4 we evaluate the prediction of four binary clinical deficits, where the blue lines indicate the AUC for *FI*_*GCP*_ vs the global cutpoint *X*_*GCP*_. The corresponding AUC of the published FI-Lab (Blodgett et al. 2017) is indicated by the horizontal orange lines. We see that *FI*_*GCP*_ is as good as FI-Lab for a range of cutpoints – approximately where mortality prediction also performs best. (All clinical deficits are tested in supplemental Fig. S8 for NHANES and Fig. S9 for CSHA.).

**Fig. 4.**
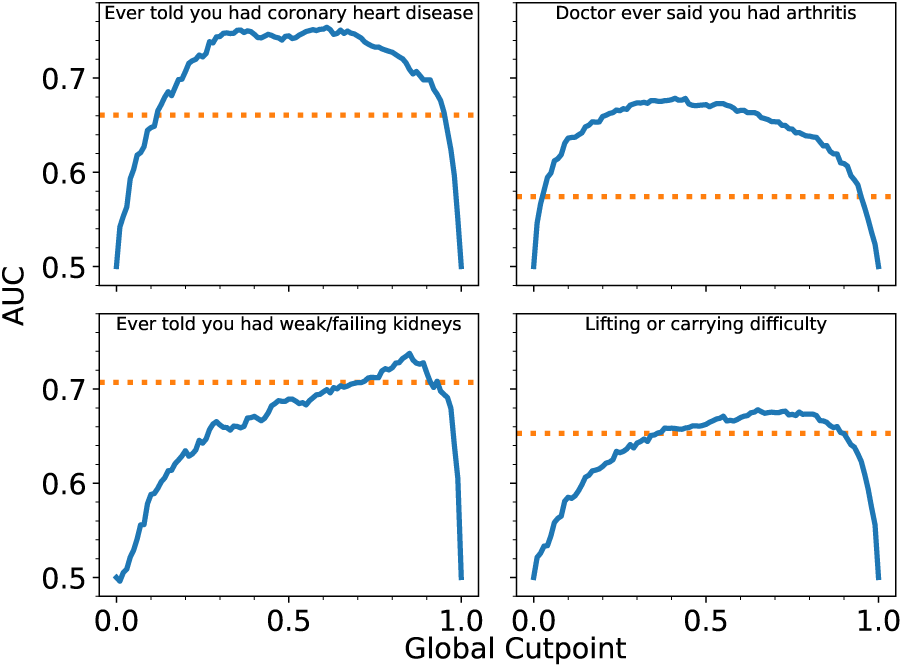
The blue lines indicate AUC of *FI*_*GCP*_ vs the global cutpoint *X*_*GCP*_ for four clinically observable deficits in the NHANES study. The horizontal dashed orange lines indicate the AUC from published FI-Lab (Blodgett et al. 2017). *FI*_*GCP*_ performs at least as well as FI-Lab, although the range of cutpoints which are most effective varies. Similar plots for all clinical deficits are shown in supplemental Fig. S8 for NHANES and Fig. S9 for CSHA.

We illustrate the distribution of *FI*_*GCP*_ in supplemental Fig. S10 for *X*_*GCP*_ = 0.85 and for *X*_*GCP*_ = 0.4. Both perform as well as FI-Lab in terms of predicting mortality (see Fig. 2). However they have very different distributions when using the same NHANES population. While *X*_*GCP*_ = 0.85 has a similar distribution as the published FI-Lab, *X*_*GCP*_ = 0.4 leads to significantly higher FI values. While this is not unexpected, since the extreme value of *X*_*GCP*_ = 0.0 would lead to all FI being equal to 1, it does lead us to examine the upper and lower limits of FI. In Fig. 5 we show the upper (light blue) and lower (dark blue) 1% of the *FI*_*GCP*_ distributions in the NHANES dataset vs *X*_*GCP*_. We see that as *X*_*GCP*_ increases both the maximum and the minimum *FI*_*GCP*_ decrease. For *X*_*GCP*_ ∼ 0.7 the minimum is zero. For *X*_*GCP*_ = 0.85 the range of maximal *FI* approximately corresponds to the range observed for the published FI-Lab (indicated in red, and labeled “Blodgett”). We also show that the 1^*st*^ and 99^*th*^ percentiles of *FI*_*GCP*_ in the CSHA dataset (black dashed lines) are similar to those of the NHANES dataset, despite the large differences in, e.g., the age distribution between these cohorts.

**Fig. 5.**
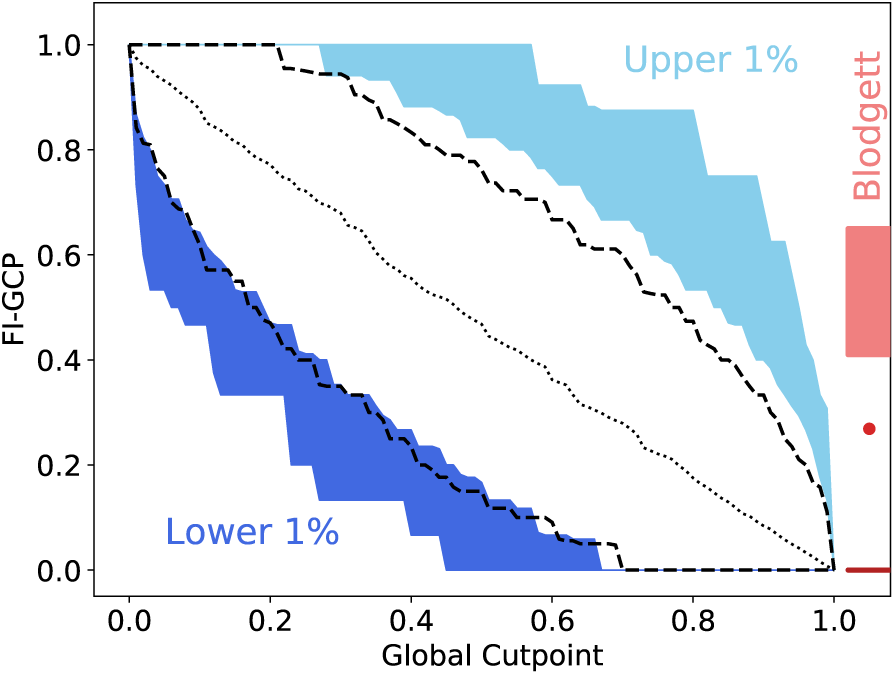
The upper 1% (light blue) and lower 1% (dark blue) of *FI*_*GCP*_ vs the global cutpoint *X*_*GCP*_ for the NHANES dataset. The dashed black lines show the 1^*st*^ and 99^*th*^ percentiles of *FI*_*GCP*_ in the CSHA dataset. The dotted diagonal black line shows the average *FI*_*GCP*_. The ranges and average for the published FI-Lab are indicated in red (Blodgett et al. 2017).

## Discussion

For a large range of global cutpoints we have shown *FI*_*GCP*_ to predict mortality and adverse clinical outcomes as well or better than FI-Lab created using established clinical risk thresholds. This result was replicated in the NHANES and CSHA data sets. Furthermore, *FI*_*GCP*_ was as informative in cross-validation, where cutpoints were calculated in one cohort and tested on another. Indeed, even applying cutpoints calculated in one age group to a cohort 20 to 40 years older remained effective. These results show *FI*_*GCP*_ is an effective method for generating an FI from biomarkers without prior knowledge of cutpoints, at least for cohorts of thousands of individuals or more.

The FI created using optimal cutpoints for each biomarker, *FI*_*logrank*_ and *FI*_*info*_, although highly informative, did not fare as well in cross-validation. Using these methods in one cohort did not yield an FI which was equivalently predictive in another cohort. Both the logrank and maximum-information based cutpoints strongly depend on the mortality of the particular cohort used and, as a result, do not represent general risk thresholds. Without extensive cross-validation we suggest these approaches be avoided.

An important question to address when implementing *FI*_*GCP*_ is which global cutpoint is appropriate. We suggest that the cutpoint be selected such that the FI has good predictive value with respect to both health outcomes and mortality. However, in both the NHANES and CSHA data-sets there is a large range of cutpoints which are similarly predictive across many of these measures. Close study of Fig. 2 indicates that *X*_*GCP*_ of 0.6 or 0.7 would build *FI*_*GCP*_ that best predicts mortality, though this range of optimal *X*_*GCP*_ may depend on the cohort. Indeed, when we consider which *X*_*GCP*_ best predicts clinical deficits, the ranges of optimal cutpoints vary significantly (see supplemental Figs. S8 and S9, particularly). It appears that there is no one “best” global cutpoint for general prediction of health outcomes, or that applies equally well across cohorts.

Another criterion for picking the global cutpoint is the interpretability of the FI within and across studies. Within the range of cutpoints which are highly predictive there are large differences in the distributions of *FI*_*GCP*_. Changing how the FI is constructed changes how individual values of the FI are assessed. For example, an FI of 0.2 has very different meaning depending on how biomarkers are binarized (see supplemental Fig. S10).

In the context of current FI studies, an appropriate global cutpoint appears to be *X*_*GCP*_ = 0.85. The resulting *FI*_0.85_ is highly predictive of both mortality and many of the clinical outcomes. Furthermore, the maximum, minimum, and mean of *FI*_0.85_ are similar to the previously published medical threshold FI-Lab. As a result, individual values of *FI*_0.85_ can be more easily interpreted between studies.

*FI*_*GCP*_ also provides a framework for investigating many aspects of the FI. Indeed, we find that some common characteristics of the FI are not generally applicable. One of the results of changing *X*_*GCP*_ is the systematic change of the extremely high (or low) FI observed in a population, as shown in Fig. 5. Variations of the maximum FI has been observed in FI-Clin (Searle et al. 2008, Gu et al. 2009, Bennett et al. 2013, Hubbard et al. 2015, Armstrong et al. 2015), FI-Lab (Blodgett et al. 2017, Howlett et al. 2014), between SHARE and SAGE multi-nation studies (Harttgen et al. 2013), in FI assembled from electronic health records (Clegg et al. 2016) or primary care data (Drubbel et al. 2013), and were found to be necessary in network models of the FI (Farrell et al. 2016). We have shown that any explicit choice of binarization changes the observed *FI*_*GCP*_ limits. Indeed, any evaluation of binarized deficits – whether biomarker or clinical – should have similar effects. Because of the broad AUC maximum with respect to *X*_*GCP*_ we have shown that such variations of the FI-max do not imply that the quality of predictions of mortality or adverse health should be adversely affected. While cohort effects contribute to observed differences of FI-max between studies, we suggest that binarization effects may dominate. In Fig. 5, the difference between the upper and lower 1% of *FI*_*GCP*_ between the NHANES and CSHA cohorts is less than when *X*_*GCP*_ is changed by only 0.1.

How might we compare FI that use different binarization approaches within the same cohort? Perhaps we shouldn’t: since the ability of FI to predict various clinical outcomes sometimes improves and sometimes degrades as *X*_*GCP*_ is changed, we can’t expect one FI to behave exactly like another. However, qualitative comparisons may be possible with reference to extremal values of FI such as shown in Fig. 5. For quantile cutpoints, we also have a formal relationship between the global cutpoint and the population average of the FI that should facilitate such qualitative comparisons:

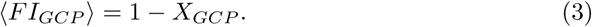

This follows since it is precisely the fraction 1 − *X*_*GCP*_ of the biomarkers which are labelled at risk, across all biomarkers. This relationship is shown as a dashed black line in Fig. 5 and appears to hold approximately for the published NHANES data as well. This remains to be better explored in future work.

Cohort effects become evident when the same cutpoint approaches are used between studies. While *FI*_*GCP*_ behaved qualitatively similarly in the NHANES and CSHA cohorts, it does show significant quantitative differences (see e.g. Fig. 2) – indicating cohort effects. *FI*_*GCP*_ is convenient for exploring cohort effects since it allows a complete separation of the cohorts at the level of biomarker binarization. For example, in previous work on FI-Lab in the CSHA and NHANES studies (Blodgett et al. 2017, Mitnitski et al. 2015) some cutpoints were sex specific (blood pressure, creatinine, blood urea, and hemoglobin) and some were not. Using *FI*_*GCP*_ we could treat all biomarkers in a generic sex specific manner by first separating the population by sex then calculating the rank normalized scores. This approach does not require previous knowledge of the cohort dependence of the biomarkers, and should be useful in future studies of general cohort dependence of the FI – including sex differences.

More generally, we have shown that FI created using population-based approaches can effectively treat biomarkers without prior medical knowledge. The same data-based approaches could also be useful in approaching FI for metabolomics, proteomics, and other omics-style applications. There is no Henry’s clinicians handbook (McPherson 2017) to select omics cutpoints from, and the large number of measurements in an omics dataset necessitates an automated method for treating potential deficits. An FI based on omics data (FI-omics) would provide insight into how frailty manifests itself on the most fundamental levels, and a quantile approach should facilitate FI-omics.

Similarly, a general method of creating the FI from biomarker measurements opens the door to many more animal model applications. Previous work has been done to create an FI-Lab in laboratory mice (Kane et al. 2019). Since there is no clinical guide for treating mice, cutpoints were selected in reference to measurements in young mice. This requirement of having a healthy cohort to use as a benchmark is incompatible with studies where there is no clearly defined healthy group available. A generic approach which can be applied to any set of biomarker measurements allows the FI to be used more generally, and should then facilitate comparisons of health and aging between organismal models and human studies.

We have explored an effective quantile-based method of creating FI from biomarker data. These methods are applicable to any set of continuous valued data where information on the age or mortality of the population is available. Our results support previous approaches using quantile-based cutpoints, including allostatic load (Juster et al. 2010) and the frailty phenotype (Fried et al. 2001). The use of global quantile cutpoints minimizes the effects of overfitting and leads to similarly predictive FI for both health and mortality over a large range of cutpoints. We found that a global cutpoint *X*_*GCP*_ = 0.85 recapitulates earlier FI-Lab results. Nevertheless there is no unique “best” global cutpoint, and the chosen cutpoint affects properties of the FI such as the maximum FI or the shape of the distribution of FI over the population.

## Data Availability

All data are publicly available.

## Acknowledgements

We thank Olga Theou for stimulating discussions.

## Supplemental

### Optimal cutpoints

Optimal cutpoints are calculated by testing binarization at every possible cutpoint in the data for every biomarker with respect to mortality at 5 years.

Mutual information can be calculated following (Farrell et al. 2016). The information entropy with respect to binary mortality *M* ∈ {0, 1} is calculated as

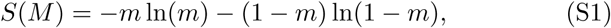

where *m* is the proportion of the population dead at 5 years. The information entropy conditional on the presence of a deficit is the average of the entropy conditioned on each state of the deficit:

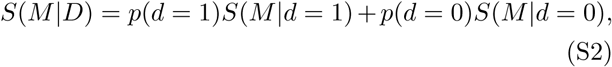

where *p*(*d*) is the proportion of the population with (*d* = 1) or without (*d* = 0) the deficit. The mutual information gained by knowing the status of a given deficit is then

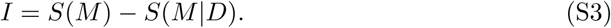

For the logrank cutpoints we use a Python implementation of the logrank test (Mantel 1966) from the survival analysis package Lifelines (Davidson-Pilon et al. 2019). Since its use of *χ*^2^ statistics for estimating p-values underestimates them systematically for small sample sizes, the logrank test has a bias to select cutpoints with extremely few individuals in one group so as to artificially decrease p-values. To compensate for this bias, we imposed a minimum group size of 20 individuals.

---

**Fig. S1.**
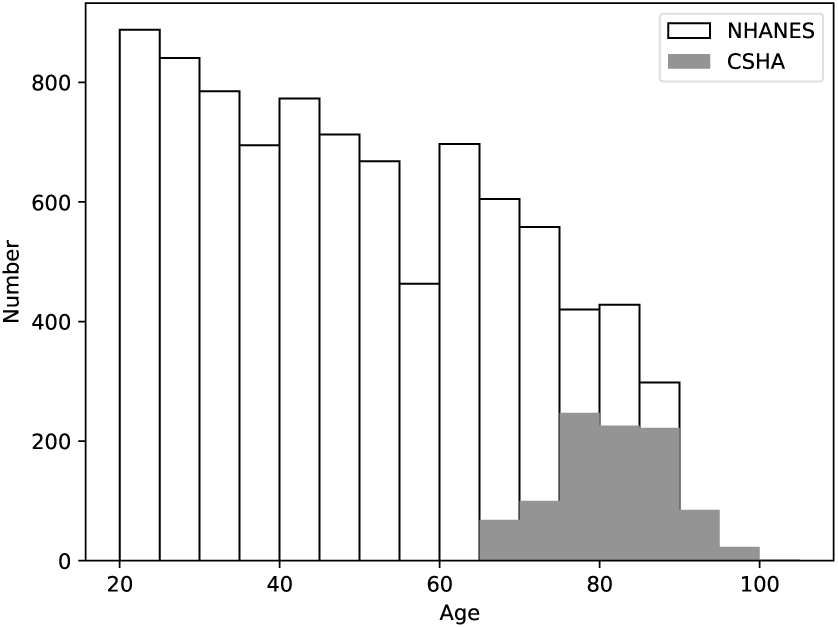
Age distributions of the CSHA (Canadian Study of Health and Aging Working Group 1994) (grey fill) and NHANES (Centers for Disease Control and Prevention National Center for Health Statistics Updated 2014) (no fill) data sets. The NHANES data set has 8881 individuals with an age range of 20 to 85 years, and is considerably larger than the CSHA study with 973 individuals. The CSHA study was limited to older individuals with ages from 65-104.

**Fig. S2.**
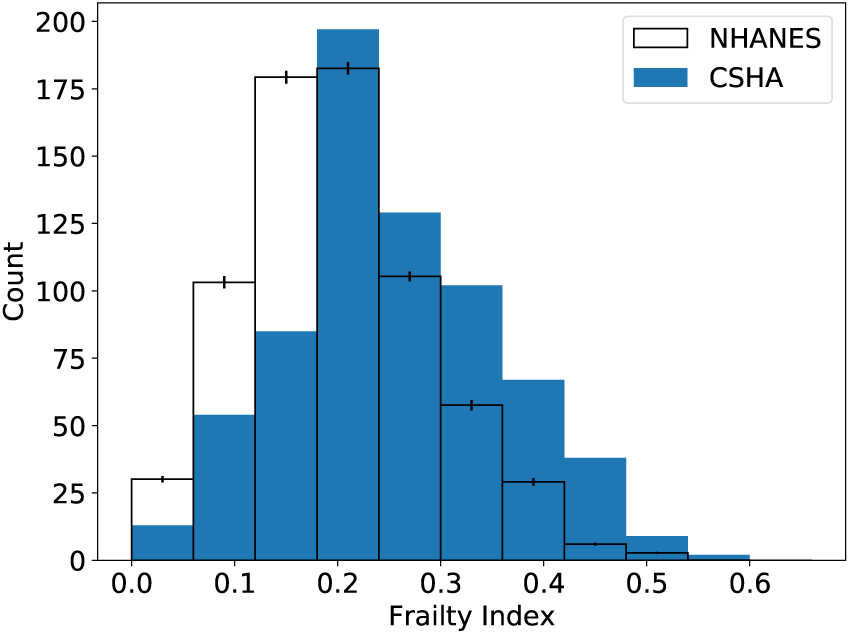
FI distributions of individuals between the ages of 65 to 85. The NHANES data (Blodgett et al. (2017)) has been randomly resampled to have the same age distribution as the CSHA data set within this age range (Howlett et al. (2014)).

**Fig. S3.**
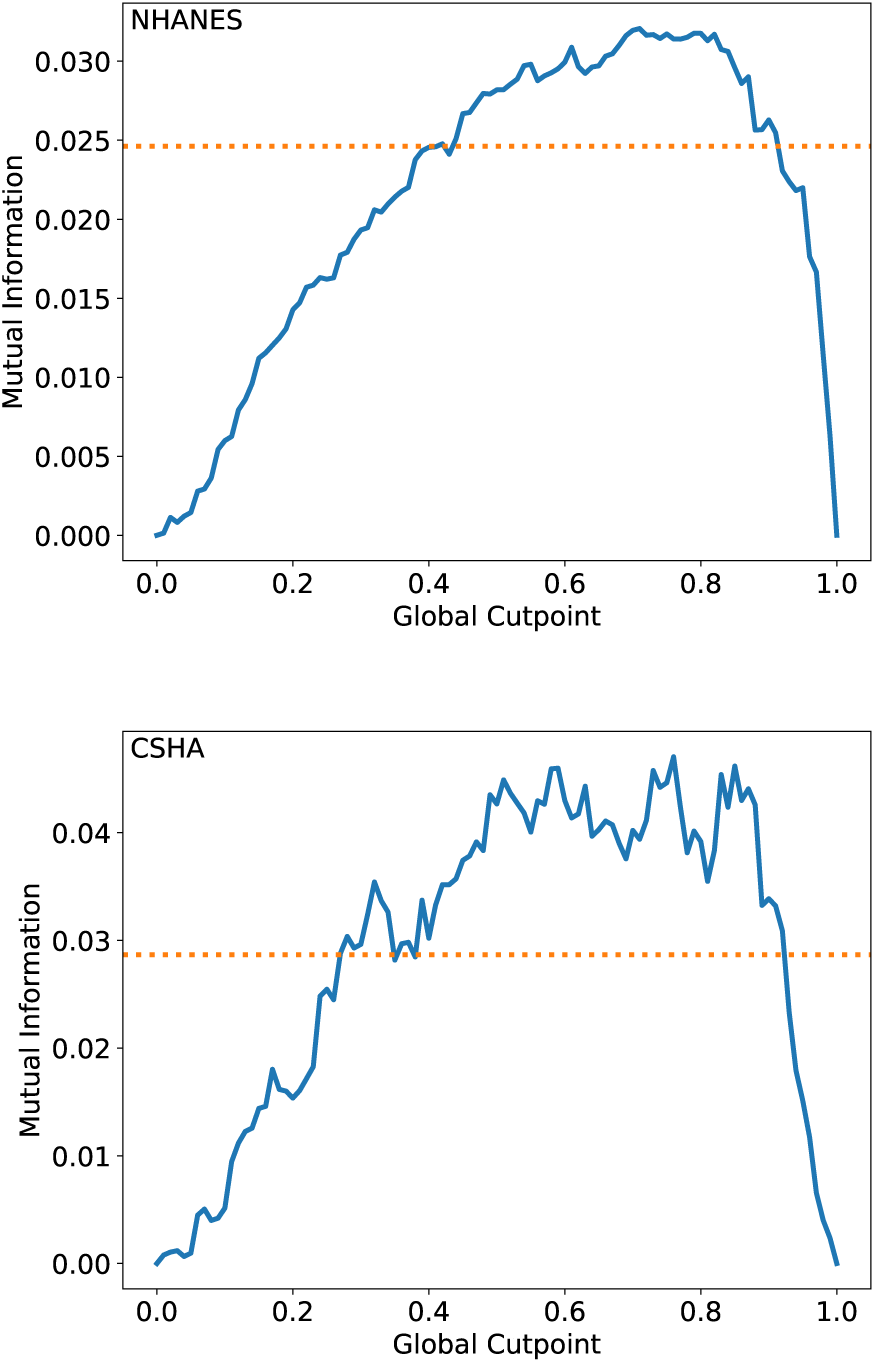
Mutual information with respect to mortality at 5 years in the NHANES (top) and CSHA (bottom) datasets for *FI*_*GCP*_ (blue lines) vs the global cutpoint *X*_*GCP*_. The orange dashed lines show the mutual information of the published FI-Lab (Blodgett et al. 2017, Howlett et al. 2014). The behavior is qualitatively like that of AUC in Fig. 2.

**Fig. S4.**
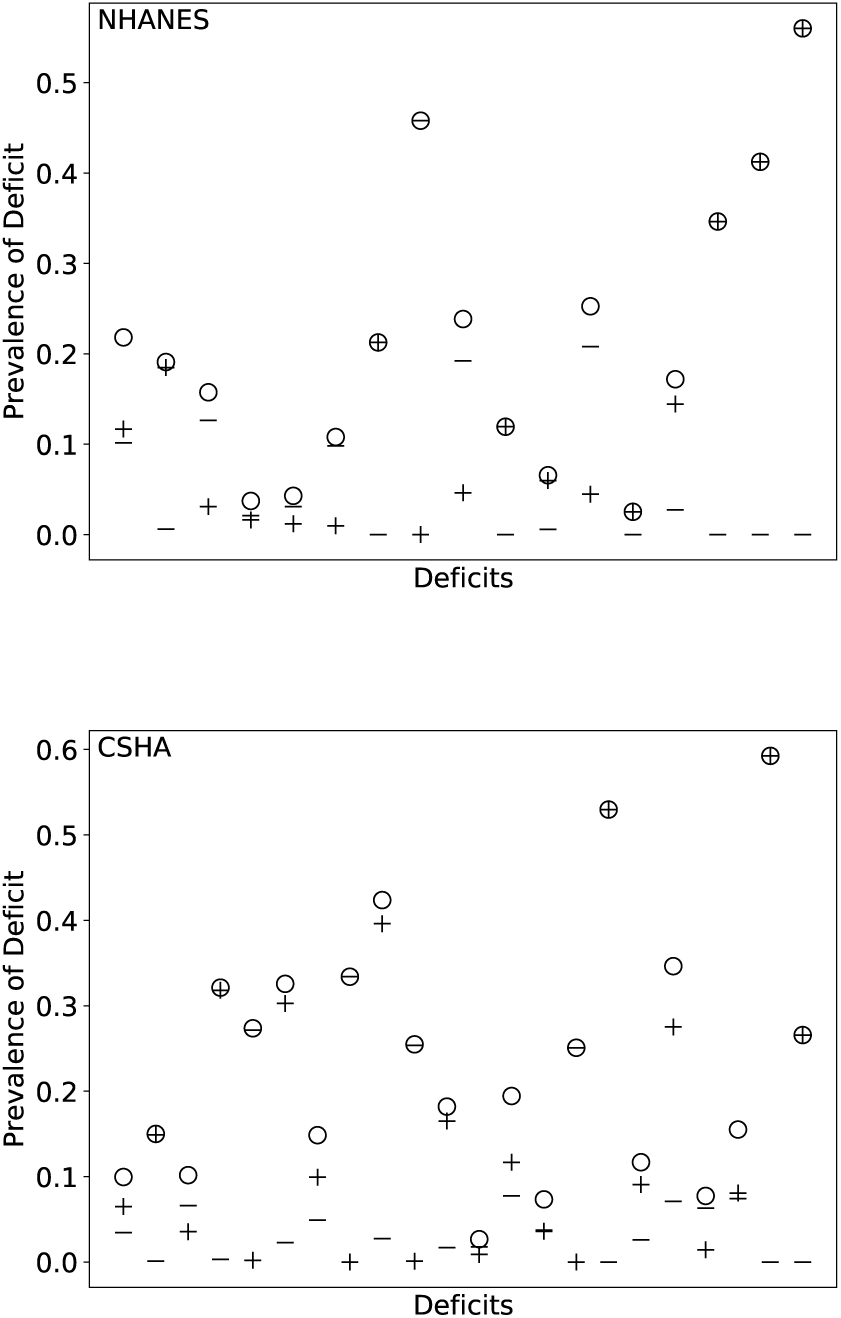
Deficit prevalence for each deficit included in the published FI-Lab (circles) broken down into proportion at risk in high (+) and low (-) categories. Most of the deficits are predominantly on a single side of the risk direction.

**Fig. S5.**
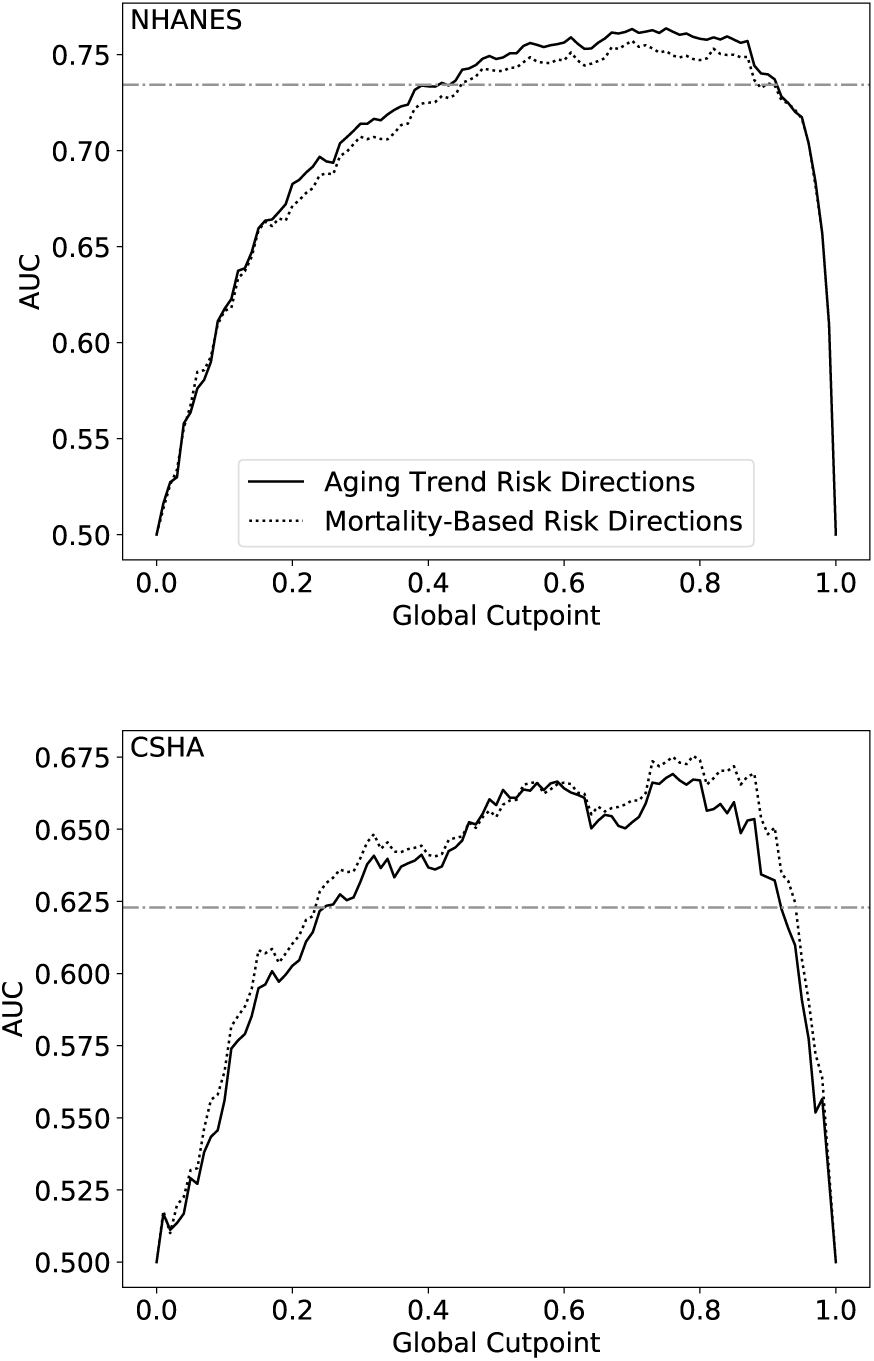
Comparing the differences in predictive value of the FI using directions of primary risk calculated with respect to mortality (dotted line) and the aging trend method (solid line) for NHANES (top) and CSHA (bottom). Note, for NHANES the age conditions are calculated only in individuals age 35 or greater, while predictive value includes the whole population. The AUC of the published FI (horizontal dot-dashed line) is provided as a benchmark.

**Fig. S6.**
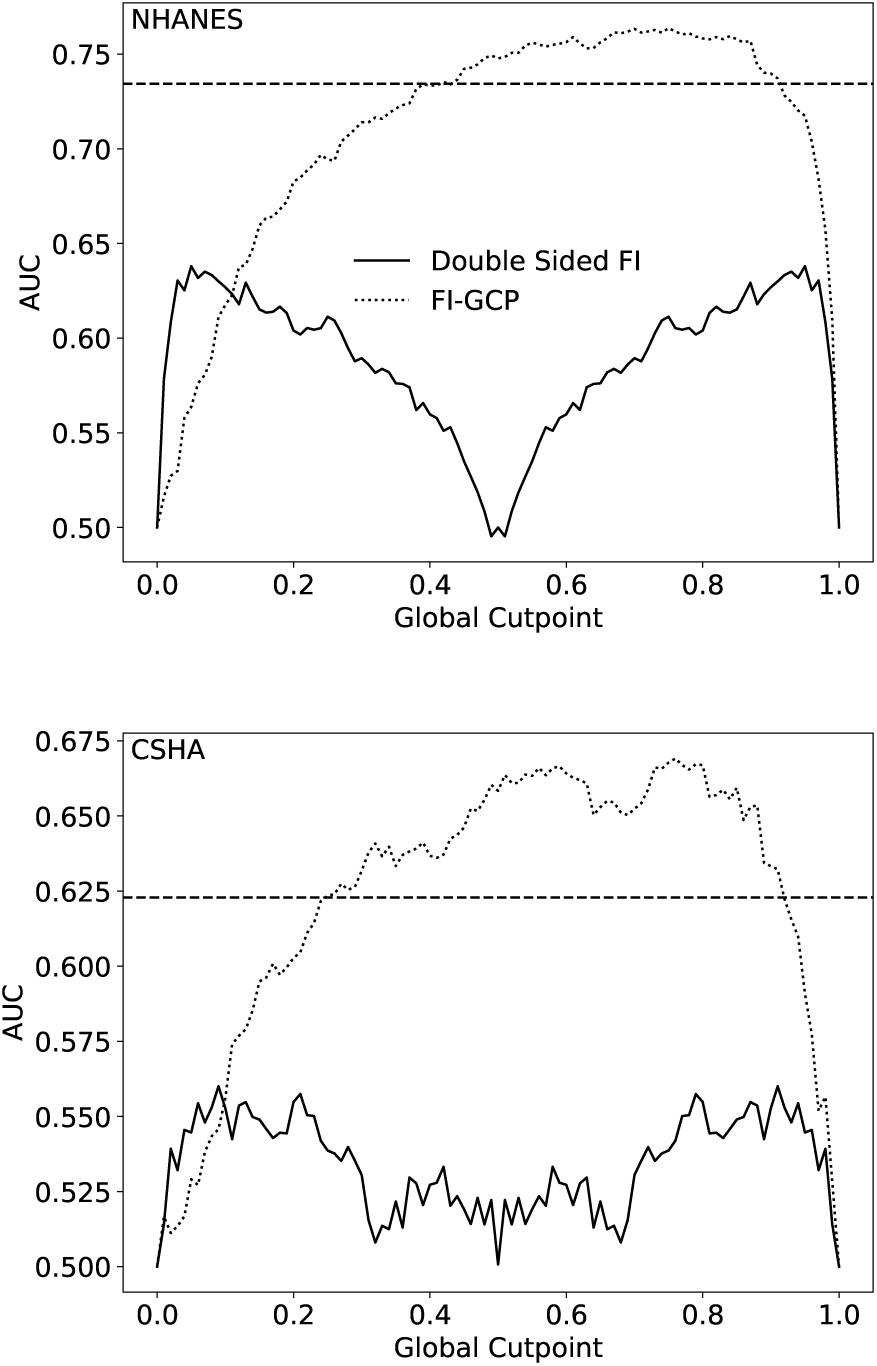
The predictive value of a symmetric two-sided binarization approach (solid line), cutpoints move out from 0.5 symmetrically. The dotted line shows the AUC of *FI*_*GCP*_, while the horizontal dashed line shows the AUC of the published FI-Lab.

**Fig. S7.**
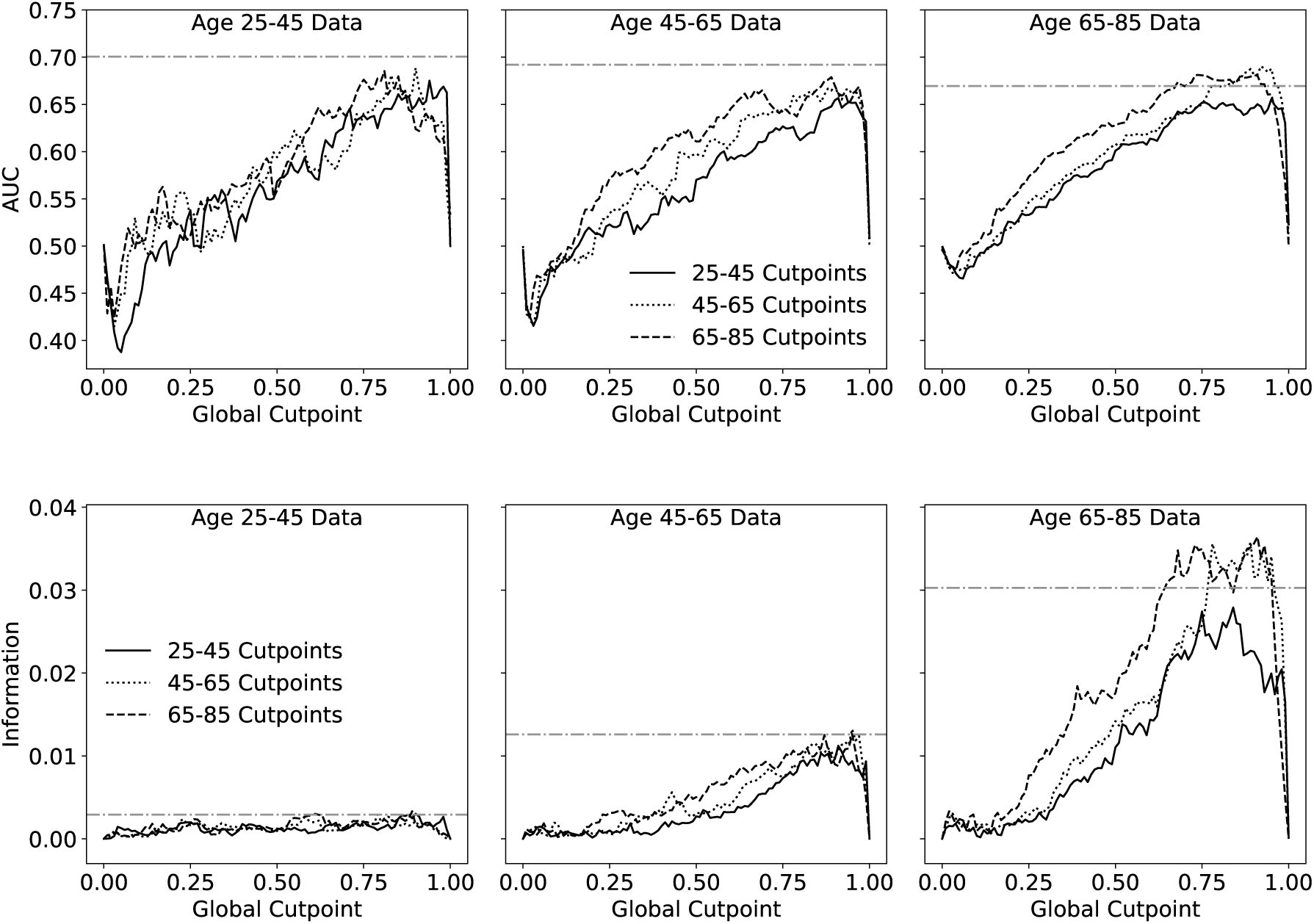
Predictive value of *FI*_*GCP*_ when cutpoints are calculated in one age group and used in another. AUC with respect to 5 year mortality is shown on top. The bottom plot shows information with respect to mortality at 5 years. Note that the information captures the poor predictive value of any FI for the youngest age group, which has very few mortality events, while the AUC does not.

**Fig. S8.**
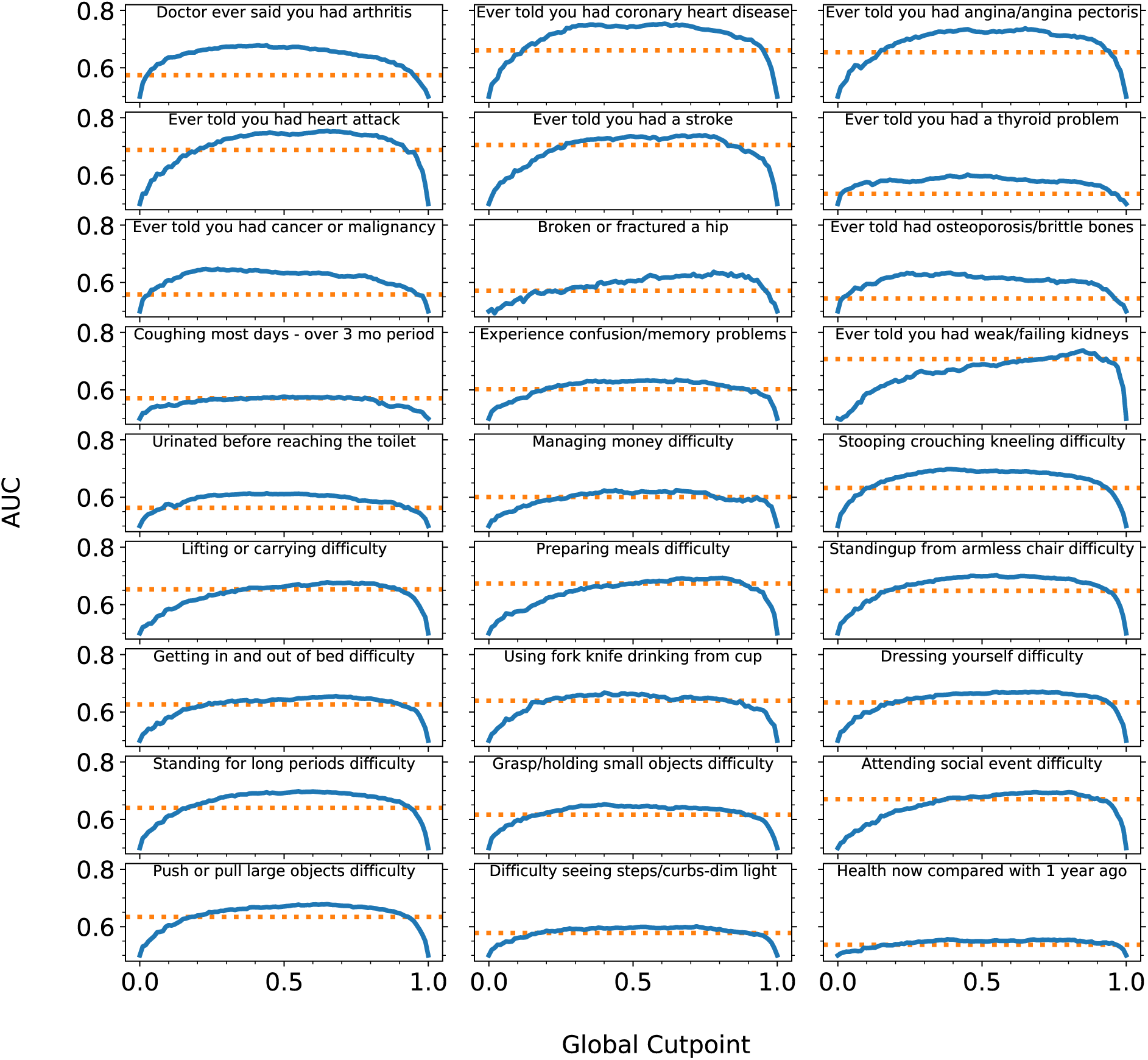
Using *FI*_*GCP*_ to “predict” clinical deficits (solid blue lines) is at least as effective as using the published FI-Lab (horizontal dotted lines) for all deficits in the NHANES study.

**Fig. S9.**
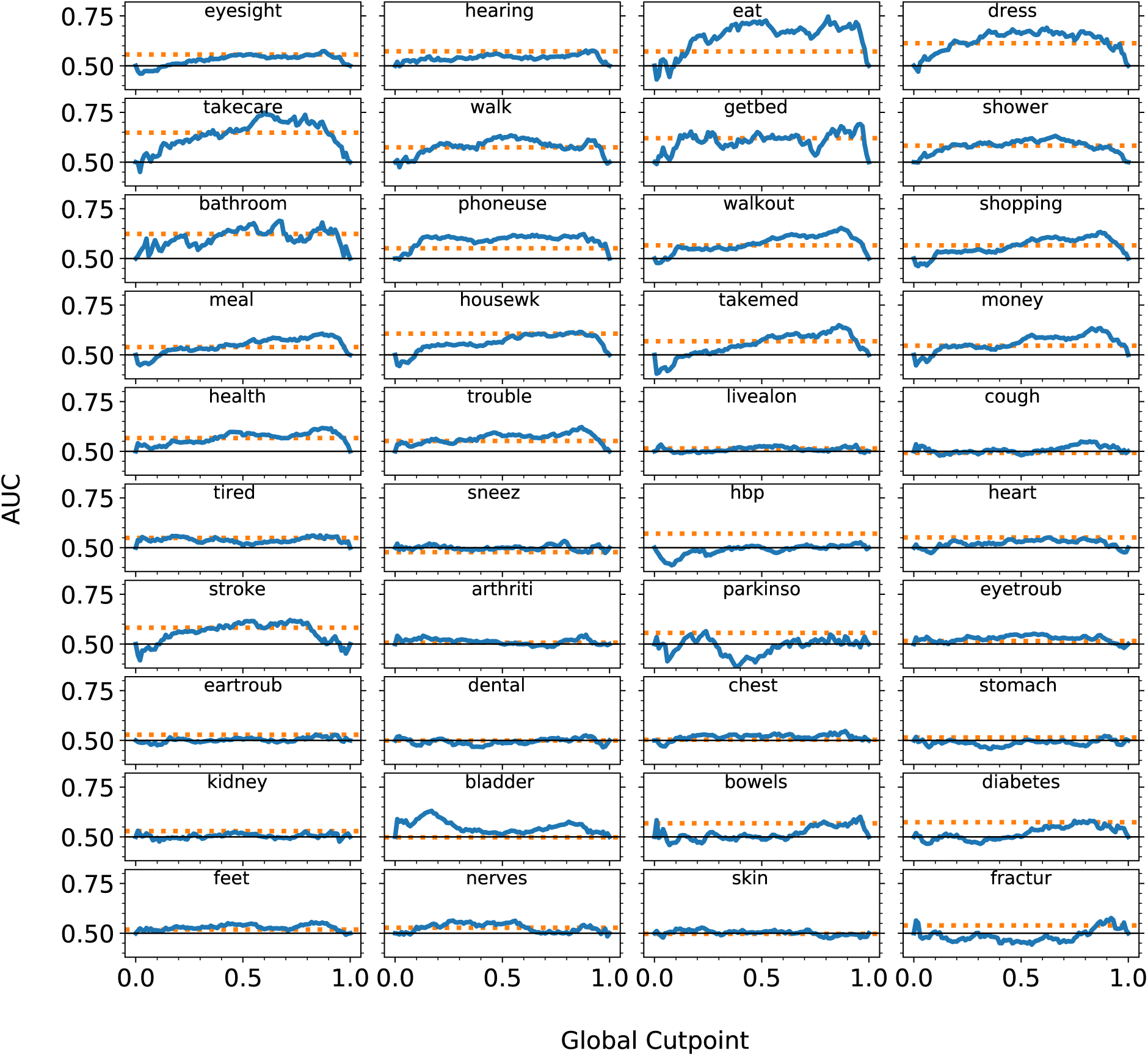
Using *FI*_*GCP*_ to “predict” clinical deficits (solid blue lines) is at least as effective as using the published FI-Lab (horizontal dotted lines) for most deficits in the CSHA study. The horizontal black line shows the benchmark AUC of 0.5 for visual reference.

**Fig. S10.**
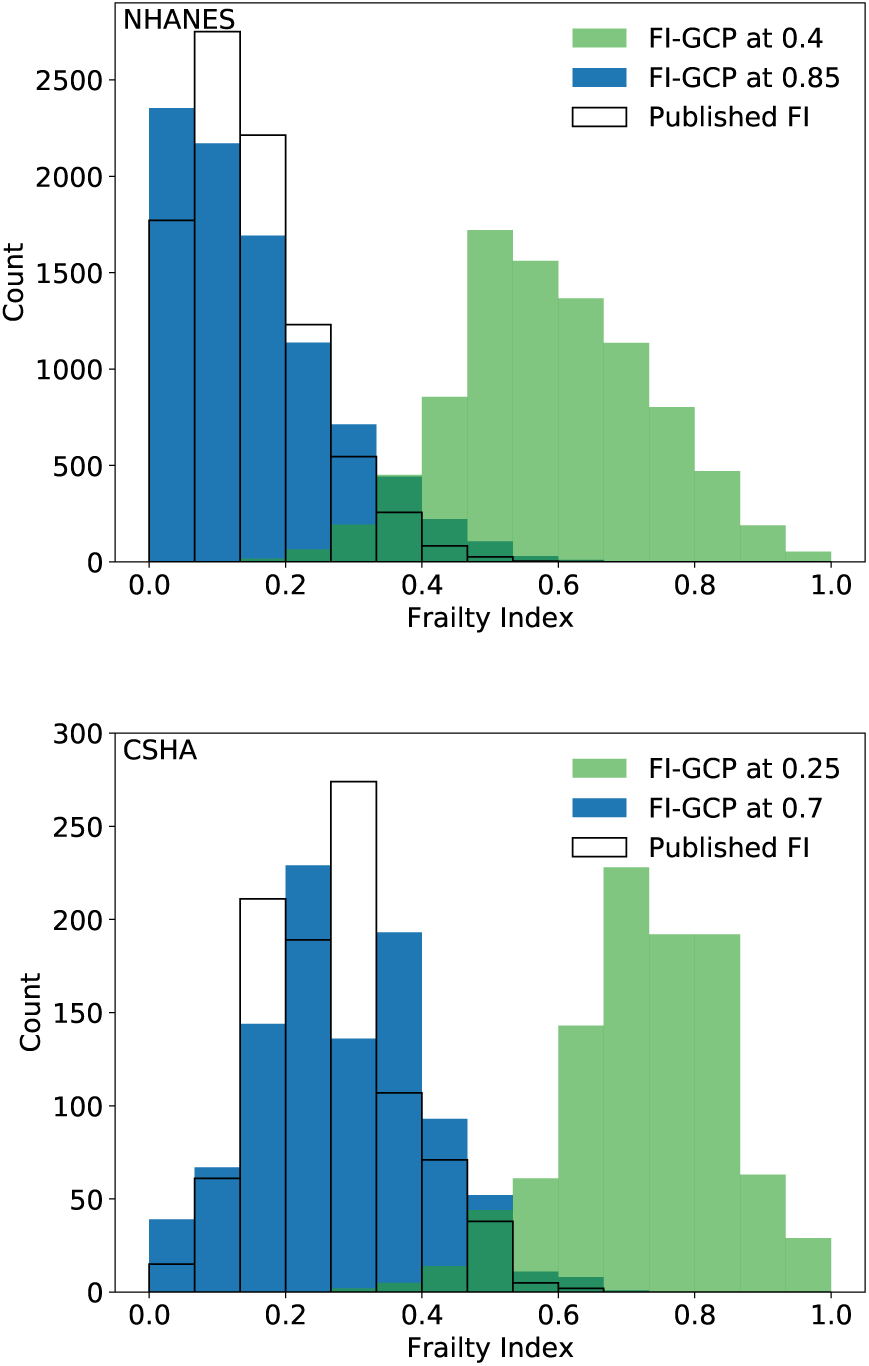
FI distributions in the NHANES (top) and CSHA (bottom) using the published FI (no fill), *FI*_*GCP*_ with cut-point at the minimum cutpoint with similar prediction to the published FI (0.4 and 0.25, for NHANES and CSHA respectively, in green), and *FI*_*GCP*_ with cutpoint where the distributions are most similar to the published FI (0.85 and 0.7, for NHANES and CSHA respectively, in blue).

